# Genome-wide association analysis identifies ancestry-specific genetic variation associated with medication response in the Study to Understand the Genetics of the Acute Response to Metformin and Glipizide in Humans (SUGAR-MGH)

**DOI:** 10.1101/2022.01.24.22269036

**Authors:** Josephine H. Li, Laura N. Brenner, Varinderpal Kaur, Katherine Figueroa, Miriam S. Udler, Aaron Leong, MAGIC Investigators, Josep M. Mercader, Jose C. Florez

**Affiliations:** Center for Genomic Medicine, Massachusetts General Hospital, Boston, Massachusetts; Diabetes Unit, Massachusetts General Hospital, Boston, Massachusetts; Programs in Metabolism and Medical & Population Genetics, Broad Institute of Harvard and MIT, Cambridge, Massachusetts; Harvard Medical School, Boston, Massachusetts; Division of Pulmonary and Critical Care Medicine, Massachusetts General Hospital, Boston, Massachusetts; Division of General Internal Medicine, Massachusetts General Hospital, Boston, Massachusetts

## Abstract

**Background:** Characterization of genetic variation that influences response to glucose-lowering medications is instrumental to precision medicine for treatment of type 2 diabetes (T2D). SUGAR-MGH examined the acute response to two anti-diabetes medications in order to understand the functional relevance of known T2D- and glycaemic trait-associated genetic loci.

**Methods:** 1,000 participants at risk for T2D from diverse ancestries underwent sequential glipizide and metformin challenges. A genome-wide association study was performed using the Illumina Multi-Ethnic Genotyping Array. Imputation was performed with the TOPMed reference panel. Multiple linear regression using an additive model tested for association between variants and primary endpoints of drug response. We evaluated the influence of 804 unique T2D and glycaemic trait-associated variants on SUGAR-MGH outcomes and performed colocalization analyses to identify shared genetic signals.

**Findings:** Five genome-wide significant variants were associated with metformin or glipizide response. The strongest association was between an African ancestry-specific variant (minor allele frequency=0·026) at rs149403252 and lower fasting glucose following metformin, adjusted for baseline glucose (*p*=1·9×10^−9^), with a 0·94 mmol/L larger decrease in fasting glucose after metformin. We identified associations between T2D-associated variants and glycaemic response, including the T2D-protective C allele of rs703972 near *ZMIZ1* and increased levels of active GLP-1 (*p*=1·6×10^−5^), supporting the role of alterations in incretin levels in T2D pathophysiology.

**Interpretation:** We present a well phenotyped, densely genotyped, multi-ancestry resource to study gene-drug interactions, uncover novel variation associated with response to common anti-diabetes medications, and provide insight into mechanisms of action of T2D-related variation.

**Funding:** US National Institutes of Health.

## INTRODUCTION

Treatment of type 2 diabetes (T2D) currently follows a standard algorithm that begins with metformin,^1^ but involves the trial and error of additional drug regimens as the disease progresses. The choice of agent is based on several considerations, including an individual’s comorbidities, the drug’s side effect profile, and costs of the therapy, but does not include information about the molecular target of the agent or genetic factors that might predict response or development of adverse effects.^2^ The understanding of who responds best to each medicine is instrumental to furthering and optimizing care of patients with diabetes.

Large-scale genome-wide association studies (GWAS) have identified over 700 genetic loci influencing T2D risk and glycaemic traits.^3-5^ Data on how genetic variation influences response to anti-diabetes medications is starting to emerge. In individuals with established T2D, GWAS have revealed novel loci for glycaemic response to metformin.^6,7^ With respect to sulfonylureas, candidate gene studies have uncovered genetic predictors of glycaemic response^8,9^ as well as sulfonylurea-induced hypoglycaemia.^10,11^ Recently, a GWAS of sulfonylurea response identified two independent loci associated with HbA1c reduction.^12^ Since the majority of pharmacogenetic studies have been conducted in those with established T2D, a genome-wide approach evaluating the response to metformin and sulfonylureas in a population at risk for developing T2D, in whom pancreatic β-cell failure is unlikely to have occurred, has not been previously done.

Moreover, the functional relevance of many T2D and glycaemic loci is not fully understood. The mechanisms leading to the development of T2D are complex, both intrinsic and extrinsic to the β cell.^13^ For instance, an intronic variant in the transcription factor 7-like gene (*TCF7L2*) is the strongest common genetic risk factor for T2D,^14^ yet multiple mechanisms have been proposed, including reduced β-cell mass, diminished insulin secretion, and alterations in the incretin response.^15^ In SUGAR-MGH, we previously observed that an impaired incretin effect may contribute to the increased risk of T2D in carriers of the high-risk allele at *TCF7L2*.^8,16^

In this study, we applied a genome-wide approach to comprehensively identify novel genetic predictors of acute metformin and glipizide response in individuals at risk of T2D but naive to these medications. We examined the effect of known genetic variants associated with T2D and glycaemic traits across all outcomes in SUGAR-MGH to gain further insights into the mechanisms by which they confer increased risk of T2D or glycaemic dysregulation. Overall, we present and make available a resource for studying how genetic variation influences the biochemical response to two common glucose-lowering agents.

## METHODS

### Study design and participants

SUGAR-MGH is a pharmacogenetic study in which 1,000 individuals who were naive to T2D medications received a single-dose glipizide challenge and a short course of metformin.^8,17^ Figure 1 summarizes the study design, which is described in detail in the Supplementary File. The study has been registered on ClinicalTrials.gov (<u>NCT01762046</u>) and is approved by the Mass General Brigham Human Research Committee IRB.

**Figure 1.**
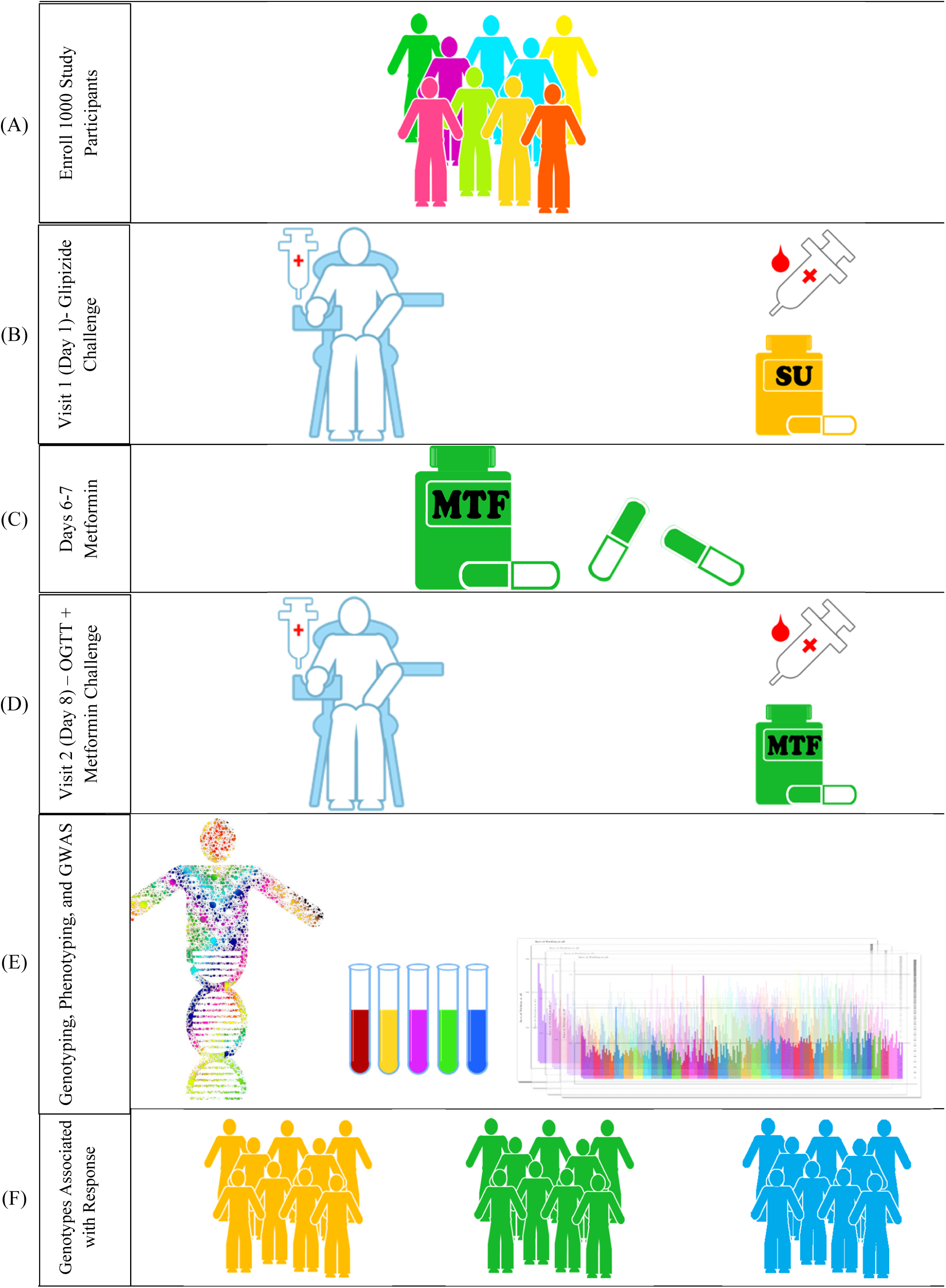
Study schema. (A) We recruited 1000 individuals at risk of developing type 2 diabetes. (B) At Visit 1, participants had their vital signs monitored, provided whole blood for DNA, and underwent fasting measurements. Individuals with a fasting blood sugar >80 mg/dL received a dose of 5 mg glipizide orally, followed by additional measurements. (C) After a 5-day wash-out period, participants received 3 doses of metformin 500 mg. (D) At Visit 2, participants returned for the final (4^th^ dose) of metformin and a 75-mg oral glucose tolerance test. (E) We performed genome-wide genotyping, constructed phenotypes of drug response, and performed a genome-wide association study, in order to (F) identify genotypes associated with outcomes of drug response.

### Genotyping and imputation

Samples underwent genome-wide genotyping on the Illumina Multi-Ethnic Genotyping Array, which covers over 1.7 million genetic markers. A three-step quality control protocol was applied using PLINK.^18^ This included two stages of variant removal and an intermediate stage of sample exclusion. Variants were filtered for minor allele frequency (MAF) <0·01, low call rate <95%, and failure to meet Hardy-Weinberg equilibrium within each self-described ancestry group (*p*<5×10^−7^). Samples were excluded for sex discrepancies, close relatedness (pairs with PI-HAT≥0·125, from which we removed the individual with the highest proportion of missingness), and call rate <98%. Phasing was performed using SHAPEIT2.^19^ Imputation was performed with the Michigan Imputation Server using the TOPMed reference panel.^20^ After post-imputation quality control, ∼12 million variants were available for analyses in 890 individuals. Genome annotations were generated using the GRCh38 assembly.

### Endpoints of metformin and glipizide response

As previously described,^17^ the primary endpoint of metformin response was defined as the fasting glucose at Visit 2 (V2), adjusted for fasting glucose at Visit 1 (V1). For the primary outcomes of glipizide response, we selected the following endpoints: insulin peak adjusted for baseline insulin, glucose trough adjusted for baseline glucose, and time to glucose trough. We identified secondary outcomes of metformin and glipizide response based on measurements taken during the glipizide challenge and the 75-g oral glucose tolerance test (OGTT) following metformin (Supplementary Table S1).

### Genome-wide association analysis

We performed genome-wide association analyses to assess the role of genetic variation in the acute response to metformin and glipizide. Multiple linear regression using an additive model tested for association between genetic variants and the primary endpoints, implemented using SNPTEST v2·5·4. Analyses were adjusted for age, sex, body mass index (BMI), and the first 10 ancestry principal components (PCs). Quantitative traits were rank-inverse normalized to avoid spurious associations driven by outliers or skewed distributions and beta estimates reflect rank-inverse normalization. When relevant, we adjusted for the baseline trait at V1. Genome-wide significance was set at *p*<5×10^−8^ and an experiment-wide threshold was set at *p*<2·5×10^−8^, accounting for two drugs. GWAS results were filtered to MAF >0·005 and imputation R^2^>0·8. Manhattan and quantile-quantile plots were produced with R (Version 4·0), and regional association plots were visualized using LocusZoom.

In an exploratory analysis, we tested for the association of variants with the pre-defined secondary drug outcomes and assessed findings that reached both genome-wide significance for at least one trait, and suggestive significance threshold of *p*<1×10^−6^ for another trait. For selected top variants of interest, we examined their association with glucose and insulin curves during the glipizide challenge and the OGTT following metformin administration. Multiple linear regression assessed for differences in outcomes by genotype groups, adjusted for similar covariates.

We also assessed the association between previously reported genome-wide significant loci for T2D and glycaemic traits and all available traits in SUGAR-MGH. We evaluated 429 genetic variants associated with T2D^3,5^ and 375 genetic variants associated with glycaemic traits.^4^ We used a r^2^ threshold of 0·5 to prune variants based on linkage disequilibrium and identify a set of independent variants, resulting in 563 effective markers. We accounted for multiple testing by setting an experiment-wide threshold of *p*<8·9×10^−5^ (0.05/563). For variants tested that met this significance threshold with any trait in SUGAR-MGH, we proceeded with colocalization analysis of the SUGAR-MGH trait and the relevant T2D/glycaemic trait to confirm the presence of shared genetic risk factors (Supplementary File).^21^

We generated weighted global extended polygenic scores (gePS) for T2D, fasting glucose, fasting insulin, and HbA1c, based on summary statistics from published GWAS of T2D and glycaemic traits.^3-5^ To construct the gePS, we used PRS-CS using auto as a global shrinkage parameter.^22^ We constructed five process-specific polygenic scores (pPS) derived from physiologically-driven clusters.^23^ We tested these scores against the primary endpoints of metformin and glipizide response and set an experiment-wide significance threshold of *p*<0·003 to account for multiple comparisons (two drugs × nine polygenic scores). We adjusted for the same covariates as in the primary GWAS.

## RESULTS

### Participant characteristics

Baseline demographics of the 890 participants with complete GWAS data are summarized (Supplementary Table S2). Approximately 53% of participants were female, the mean age was 47 years, and 37% of participants self-reported as non-white. The mean BMI was 30·2 kg/m^2^ and mean fasting glucose was 5·14 mmol/L, consistent with a population at risk for requiring future treatment of T2D. Twenty participants did not receive the glipizide challenge due to a low baseline fasting glucose and 298 terminated the challenge early for hypoglycaemia, in accordance with study protocol.

### Association of genetic variation with primary outcomes of drug response

We identified five genome-wide significant variants associated with primary endpoints of acute metformin and glipizide response, four of which met experiment-wide significance of *p*<2·5×10^−8^ (Table 1). Three variants were associated with metformin response, as measured by fasting glucose at V2, adjusted for fasting glucose at V1; two variants were associated with glipizide response, as measured by the time to glucose trough.

**Table 1.**
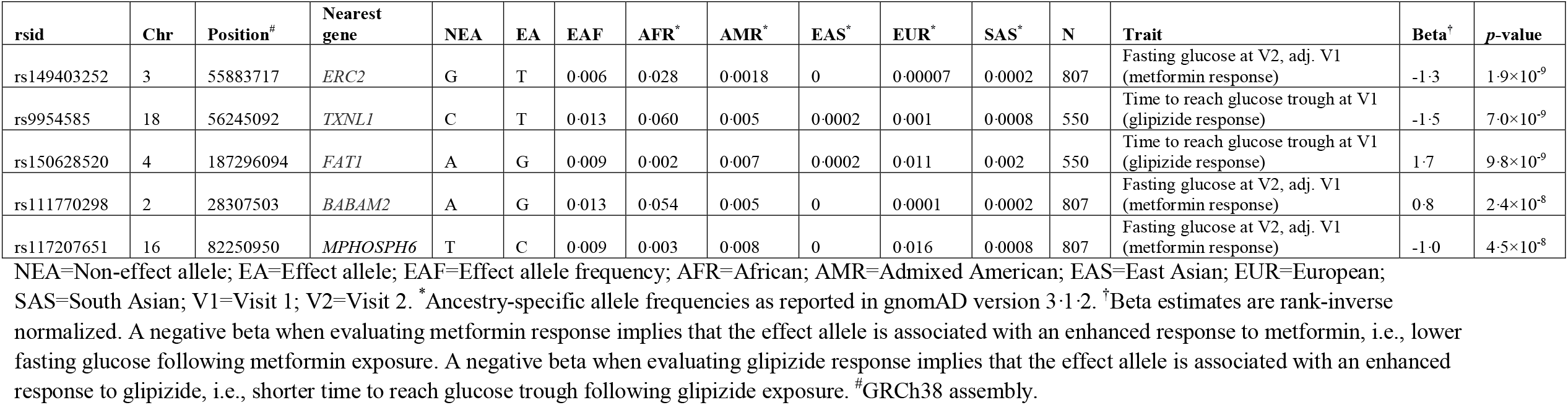
Genome-wide significant variants (*p*<5×10^−8^) associated with primary endpoints of acute metformin or glipizide response in SUGAR-MGH.

Among the variants associated with metformin response at genome-wide significance, rs149403252 (MAF(Afr)=0·039, beta=-1·3, *p*=1·9×10^−9^), is an African ancestry-specific variant located in chromosome 3 near *ERC2* (Figure 2A). Carriers of the T effect allele had a lower fasting glucose at V2, adjusted for baseline glucose, indicating that they had an enhanced metformin response. This was particularly apparent when examining the change in fasting glucose (Figure 2B), in which heterozygous individuals had a decrease of 1·1 mmol/L after four doses of metformin compared to a decrease of 0·12 mmol/L in non-carriers, (beta of difference=-0·94 mmol/L (*p*=1·1×10^−6^)). During the OGTT following metformin treatment, heterozygous individuals had lower insulin area under the curve (AUC) (*p*=0·005) despite statistically similar glucose AUC (Figures 2C-D). Another African ancestry-specific genetic variant influencing metformin response was rs111770298 (MAF(Afr)=0·053, beta=0·8, *p*=2·4×10^−8^)), located near *BABAM2* in chromosome 2 (Supplementary Figure S2). Carriers of the G allele had a reduced metformin response, as evidenced by a higher fasting glucose at V2, adjusted for baseline glucose at V1. We calculated that whereas individuals homozygous for the A (common) allele experienced a 0·15 mmol/L decrease in fasting glucose after metformin, heterozygous individuals had a 0·29 mmol/L increase (beta of difference=0·43 mmol/L (*p=*9·4×10^−7^)).

**Figure 2.**
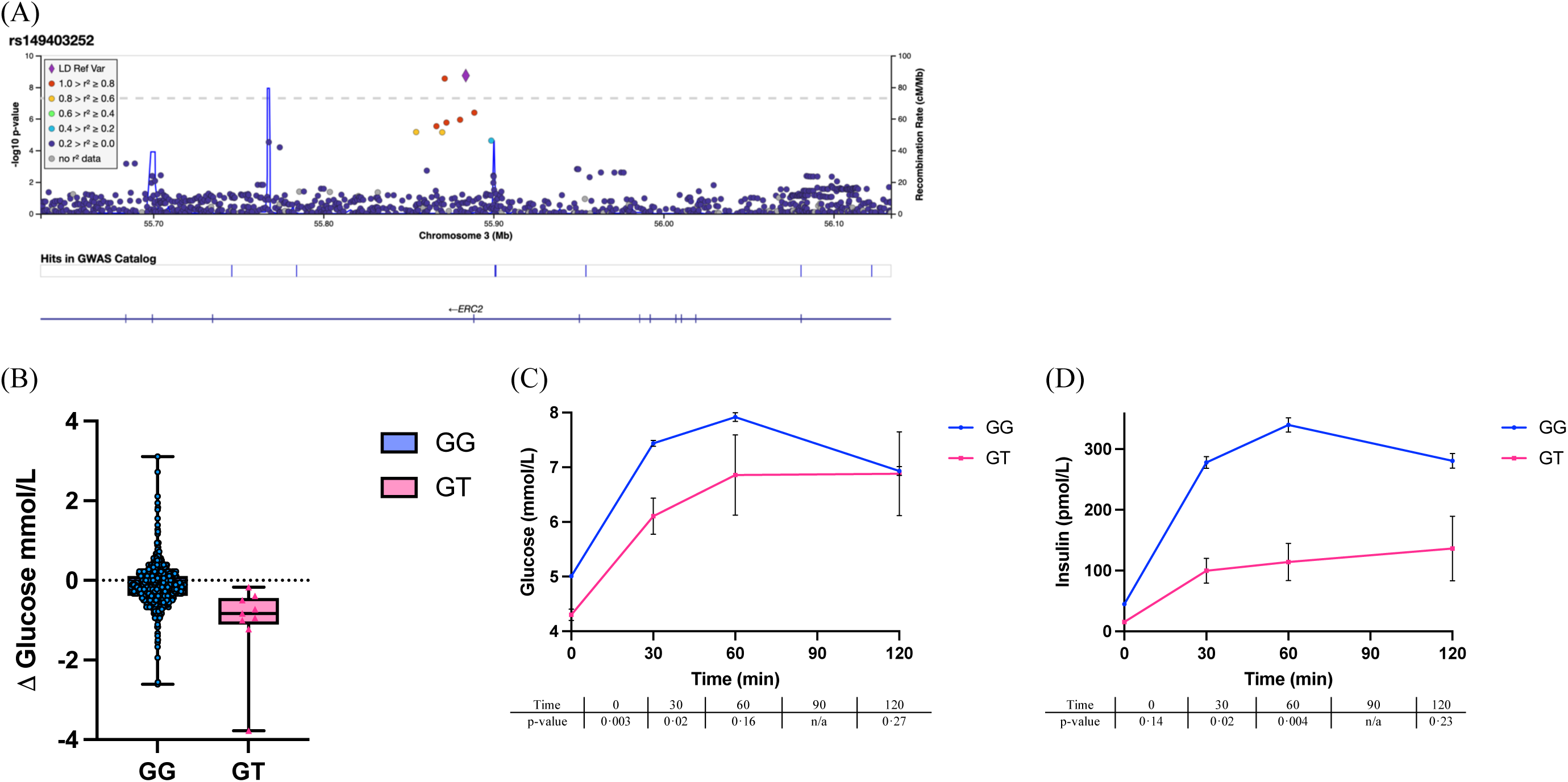
(A) Regional association plot of rs149403252. (B) Box plot illustrating mean change in fasting glucose (Visit 2 minus Visit 1) by rs149403252 genotype. (C) Change in plasma glucose by rs149403252 genotype across the OGTT following metformin. (D) Change in plasma insulin by rs34872471 genotype across the OGTT following metformin.

For the glipizide challenge, rs150628520, a low frequency variant near *FAT1* in chromosome 4 (Figure 3A), was associated with increased time to glucose trough, consistent with a diminished glipizide response (MAF=0·009, beta=1·7, *p*=9·8×10^−9^). In agreement with this, carriers of the G allele had a significantly decreased cumulative drop in glucose, measured by glucose area over the curve (Figure 3B, *p=*0·004), as well as a decreased insulin AUC (Figure 3C, *p*=0·006). Supplementary Figures S1 and S3 depict the regional association plots as well as the respective glucose and insulin curves for the remaining significant variants, located in or near *TXNL1* and *MPHOSPH6* (Table 1).

**Figure 3.**
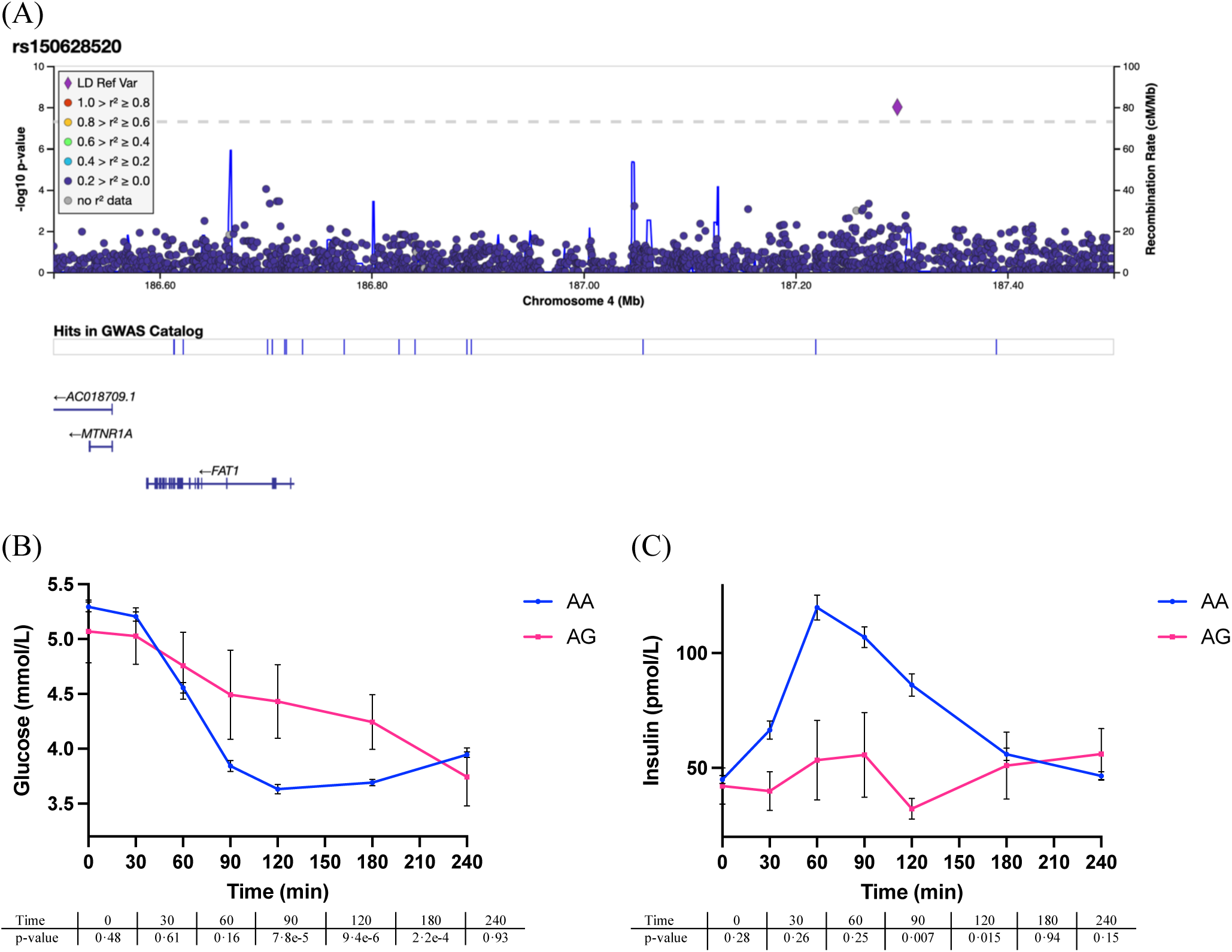
(A) Regional association plot of rs150628520. (B) Change in plasma glucose by rs150628520 genotype at Visit 1 after glipizide administration. (C) Change in plasma insulin by rs150628520 genotype at Visit 1 after glipizide administration.

### Association of genetic variation with secondary outcomes of drug response

Beyond the primary outcomes, we examined associations reaching genome-wide significance (*p*<5×10^−8^) for either the primary or secondary outcomes (Supplementary Table S1). We curated a list of variants that were additionally associated with at least one other secondary outcome at a suggestive *p*<1×10^−6^ and in consistent direction of effect (i.e., both associations pointing toward enhanced metformin response). Supplementary Table S3 describes the resultant set of markers that met that criteria and therefore considered true associations with metformin or glipizide response. Two of the primary GWAS findings (rs150628520 and rs111770298) were also associated with secondary outcomes (Supplementary Table S3). Of the remaining five variants, rs146209333 (MAF=0·012), an intronic variant in *PRICKLE2* (Supplementary Figure S4), was associated with the greatest number of secondary outcomes, namely lower glucose at 60 min (beta=-0·9, p=3·3×10^−8^), steeper slope to glucose trough (beta=1·2, p=9·9×10^−8^), and lower glucose trough (beta=-0·9, p=2·8×10^−6^). Consistent with these findings, in an independent analysis, each C allele was associated with 5·4 increased odds of terminating the glipizide challenge due to hypoglycaemia (*p*=0·002).

### Evaluation of known T2D and glycaemic variation and polygenic scores in SUGAR-MGH

We next focused on assessing the pharmacological response to variants previously associated with T2D and glycaemic traits. Within the top associations meeting experiment-wide significance threshold of *p*<8·9×10^−5^ in any of the SUGAR-MGH outcomes, we were able to confirm through colocalization analyses that 10 of them share the same genetic signal between a SUGAR-MGH outcome and T2D/glycaemic traits with a posterior probability (PP) of ≥75% (Table 2). As an example, we found that the T2D-protective C allele of rs703972 near *ZMIZ1* was associated with increased levels of active glucagon-like peptide 1 (GLP-1) (*p*=1·6×10^−5^), with high evidence of colocalization (PP=90·3%, Figure 4A-C). Moreover, as previously reported,^8^ the well-described T risk allele of rs7903146 in *TCF7L2* appeared to influence higher total and active GLP-1 levels in SUGAR-MGH and demonstrated good colocalization with both T2D (Figure 4D-F) and glycaemic traits (Table 2).

**Table 2.** Known genome-wide significant T2D and glycaemic variation meeting experiment-wide significance in SUGAR-MGH (*p*<8·9×10^−5^) and colocalization results.

| rsid | Chr | Position<sup>#</sup> | Nearest gene | NEA | EA | EAF* | OR (95% CI)* | T2D $p$-value* | | SUGAR-MGH trait | N | Beta<sup>†</sup> | $p$-value | PP result |
| --- | --- | --- | --- | --- | --- | --- | --- | --- | --- | --- | --- | --- | --- | --- |
| rs703972 | 10 | 79193069 | *ZMIZ1* | G | C | 0.467 | 0.93 (0.92-0.94) | $1.7 \times 10^{-29}$ | | Active GLP-1 at 30 min at V2 | 143 | 0.5 | $1.6 \times 10^{-5}$ | 90.3 |
| rs9828772 | 3 | 129614339 | *TMCC1* | C | G | 0.102 | 0.94 (0.93-0.96) | $4.2 \times 10^{-8}$ | | Fasting glucagon at V1 | 492 | 0.4 | $2.1 \times 10^{-5}$ | 76.8 |
| rs6070625 | 20 | 58819573 | *GNAS* | C | G | 0.517 | 1.05 (1.04-1.06) | $5.3 \times 10^{-14}$ | | Fasting glucose at V2 minus fasting glucose at V1 | 805 | 0.2 | $2.9 \times 10^{-5}$ | 96.6 |
| rs11688682 | 2 | 120590036 | *GLI2* | G | C | 0.272 | 0.95 (0.94-0.97) | $4.2 \times 10^{-9}$ | | Active GLP-1 at 5 min at V2 | 144 | 0.6 | $3.1 \times 10^{-5}$ | 70.9 |
| rs12048743 | 1 | 205145745 | *DSTYK* | C | G | 0.442 | 1.04 (1.03-1.05) | $3.5 \times 10^{-9}$ | | Fasting glucose at V2 minus fasting glucose at V1 | 805 | -0.2 | $8.2 \times 10^{-5}$ | 89.7 |
| rs7903146 | 10 | 112998590 | *TCF7L2* | C | T | 0.294 | 1.37 (1.35-1.39) | $6.0 \times 10^{-447}$ | | Total GLP-1 at 10 min at V2 | 142 | 0.4 | $8.8 \times 10^{-5}$ | 79.7 |
| rsid | Chr | Position | Nearest gene | NEA | EA | EAF* | Beta (SE)* | MAGIC $p$-value* | MAGIC trait | SUGAR-MGH trait | N | Beta | $p$-value | PP result |
| rs8914 | 11 | 46677574 | *ATG13* | G | A | 0.108 | -0.040 (0.006) | $4.1 \times 10^{-13}$ | Fasting glucose | Fasting glucose at V1 | 851 | -0.3 | $6.8 \times 10^{-6}$ | 1.9 |
| rs8914 | 11 | 46677574 | *ATG13* | G | A | 0.108 | -0.040 (0.006) | $4.1 \times 10^{-13}$ | Fasting glucose | Glucose at 30 min at V1 | 832 | -0.3 | $2.3 \times 10^{-5}$ | 1.7 |
| rs8914 | 11 | 46677574 | *ATG13* | G | A | 0.108 | -0.040 (0.006) | $4.1 \times 10^{-13}$ | Fasting glucose | Fasting glucose at V2 | 807 | -0.3 | $4.9 \times 10^{-5}$ | 1.6 |
| rs2745353 | 6 | 127131790 | *RSPO3* | C | T | 0.516 | 0.031 (0.004) | $4.6 \times 10^{-16}$ | Fasting insulin | Insulin peak at V1 | 830 | 0.2 | $7.7 \times 10^{-5}$ | 91.0 |
| rs2971669 | 7 | 44192179 | *GCK* | C | T | 0.202 | 0.083 (0.008) | $3.8 \times 10^{-28}$ | 2-hour glucose | Fasting glucose at V2 | 807 | 0.2 | $8.6 \times 10^{-5}$ | 76.8 |
| rs7903146 | 10 | 112998590 | *TCF7L2* | C | T | 0.275 | 0.047 (0.004) | $2.0 \times 10^{-35}$ | Fasting glucose | Total GLP-1 at 10 min at V2 | 142 | 0.4 | $8.8 \times 10^{-5}$ | 77.0 |
| rs7903146 | 10 | 112998590 | *TCF7L2* | C | T | 0.278 | -0.027 (0.005) | $1.2 \times 10^{-9}$ | Fasting insulin | Total GLP-1 at 10 min at V2 | 142 | 0.4 | $8.8 \times 10^{-5}$ | 74.6 |
| rs7903146 | 10 | 112998590 | *TCF7L2* | C | T | 0.281 | 0.043 (0.004) | $1.0 \times 10^{-22}$ | HbA1c | Total GLP-1 at 10 min at V2 | 142 | 0.4 | $8.8 \times 10^{-5}$ | 79.4 |
| rs7903146 | 10 | 112998590 | *TCF7L2* | C | T | 0.259 | 0.070 (0.007) | $2.8 \times 10^{-26}$ | 2-hour glucose | Total GLP-1 at 10 min at V2 | 142 | 0.4 | $8.8 \times 10^{-5}$ | 80.0 |

NEA=Non-effect allele; EA=Effect allele; EAF=Effect allele frequency; PP=posterior probability; MAGIC=Meta-Analysis of Glucose and Insulin-related traits Consortium. \*Denote results from T2D GWAS and MAGIC. <sup>†</sup>Beta estimates are rank-inverse normalized. <sup>#</sup>GRCh38 assembly.

**Figure 4.**
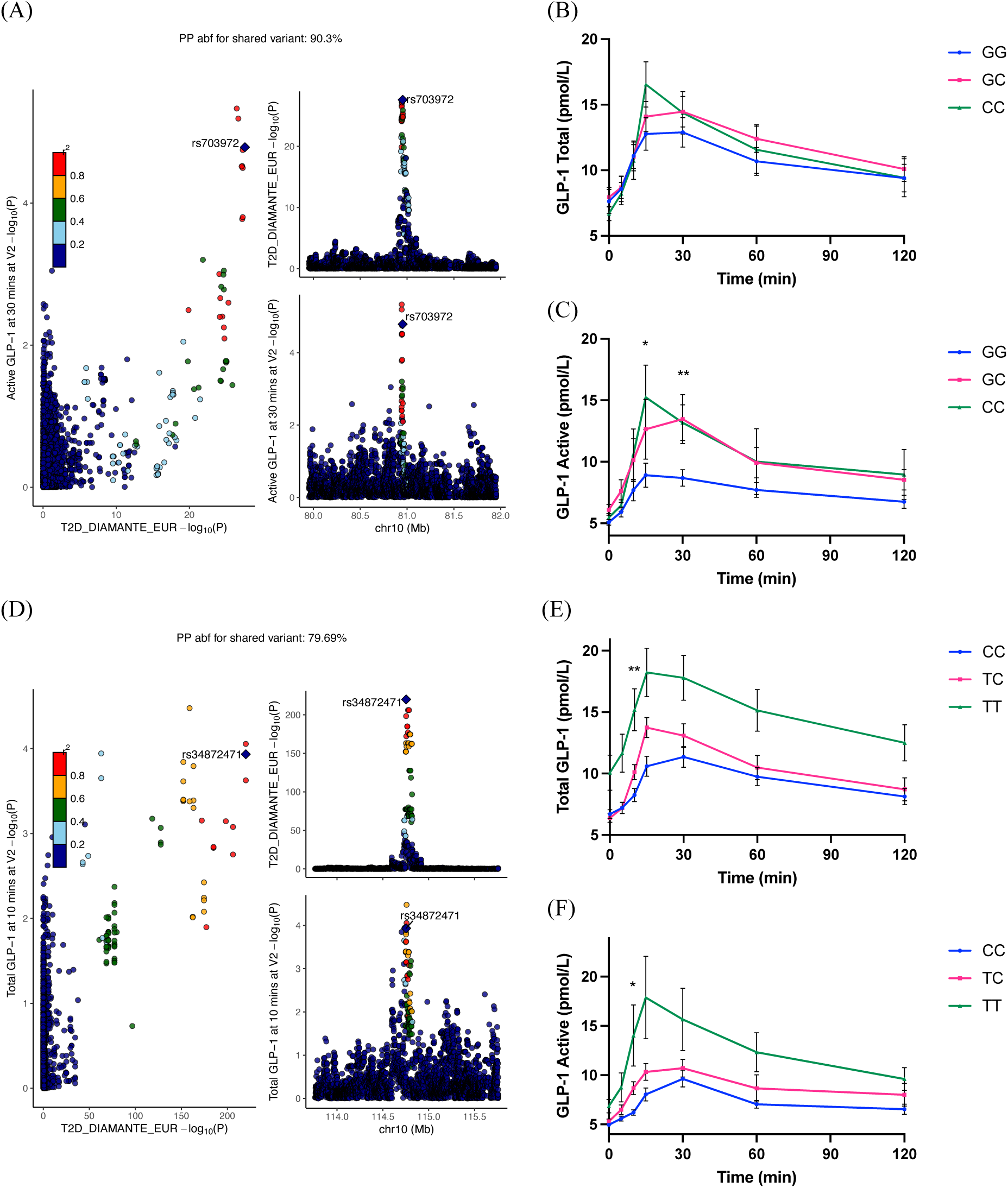
(A) Colocalization plot showing rs703972 near *ZMIZ1* colocalizes with active GLP-1 and T2D risk. LocusZoom plots of association with T2D (top right), with active GLP-1 (bottom right), and their colocalization (left plot). (B) Change in total and (C) active GLP-1 by rs703972 at V2 during oral glucose tolerance test. (D) Colocalization plot demonstrating that variants (including rs7903146 in *TCF7L2*) associated with T2D risk colocalize with total GLP-1. LocusZoom plots of association with T2D (top right), with total GLP-1 (bottom right), and their colocalization (left plot). (E) Total and (F) active GLP-1 by rs7903146 at V2 during oral glucose tolerance test. \**p*<1×10^−3^ \*\**p*<1×10^−4^

When we evaluated the influence of polygenic scores on drug response, we confirmed our previously-reported nominal association between a higher T2D polygenic score and greater glucose area over the curve (AOC), representing a greater cumulative drop in glucose following glipizide (*p*=0·02).^24^ In addition, we observed an association between the fasting glucose gePS and the primary outcome for metformin response meeting experiment-wide significance (*p=*0·002); after adjusting for baseline glucose, individuals with higher fasting glucose gePS had a 0·03 mmol/L lower drop in fasting glucose after metformin per SD increase in polygenic score, consistent with a worse metformin response. In a subgroup analysis, this was found to be driven by individuals who began the study with a fasting glucose over 5 mmol/L, who experienced a drop of 0·07 mmol/L (*p*=0·04). No associations between any of the pPS and metformin or glipizide response met experiment-wide significance (Supplementary Table S4).

## DISCUSSION

SUGAR-MGH is a pharmacogenetic resource for characterizing genetic influences on pharmacological perturbations relevant to T2D. In prior work, SUGAR-MGH has contributed to the understanding of the influence of *TCF7L2* and *CYP2C9*, as well as a restricted-to-significant (i.e., using only genome-wide significant variants) T2D polygenic risk score on drug response.^8,11,24^ With the completion of genome-wide genotyping, we report new genetic associations with acute metformin and sulfonylurea response in an ancestrally diverse population at risk for T2D and naive to commonly-prescribed anti-diabetes medications.

Our lead finding was the association of rs149403252, an intronic variant located in *ERC2*, with lower fasting glucose at V2 following metformin. The ERC2 protein is a member of the active-zone specific protein family and is also known as CAST.^25^ The CAST family contains the ELKS protein (also known as ERC1). ELKS and CAST show high amino acid homology and form oligomers with each other.^26^ These proteins have been implicated in the calcium-dependent exocytosis of neurotransmitters in the brain. ELKS has been shown to be found in pancreatic β cells and may be involved in the regulation of insulin secretion.^26^ CAST/ERC2 can be hypothesized to have a similar mechanism of exocytosis affecting insulin secretion, and an alteration in the function of ERC2 conferred by the rs149403252 variant may be responsible for the differences observed in insulin secretion and change in fasting glucose following metformin. Fine-mapping analyses, phenome-wide association analyses, and functional experiments will be needed to confirm the implication of ERC2 in metformin response.

We uncovered several promising variants of interest for glipizide response. G-allele carriers at rs150628520 near *FAT1* appear to have an attenuated response to glipizide, having not only an increased time to glucose trough but also a more gradual slope to glucose trough. Conversely, those with the C allele at rs146209333 in *PRICKLE2* have a more robust response to glipizide, with markedly lower glucose levels at multiple timepoints during the glipizide challenge and a steeper slope to glucose trough. C-allele carriers were also at higher risk of terminating the glipizide challenge early due to hypoglycaemia. *PRICKLE2* is a highly-conserved protein involved in the non-canonical Wnt and planar cell polarity signalling pathways, and pathogenic variants in *PRICKLE2* have been associated with neurodevelopmental disorders including epilepsy in humans.^27^ If this variant additionally plays a role in the pathogenesis of epileptic activity, there may be clinical implications for carriers to avoid sulfonylureas to minimize the risk of hypoglycaemia, a known risk factor for seizures. We note that the presence of concordant associations across multiple primary and secondary outcomes for glipizide response provides support for our genetic findings. However, it is unclear whether the observed differences by genotype are due to a decrease in glipizide action or impairment in glipizide absorption. To further elucidate the mechanisms responsible for these effects, future directions include quantifying glipizide drug levels, comparing carriers and non-carriers at these loci.

We also leveraged the phenotypic outcomes constructed in this physiological study to provide functional characterization of genetic variants associated with T2D and glycaemic traits. We found established genome-wide significant T2D and glycaemic variation that met experiment-wide significance for traits in SUGAR-MGH, with evidence of good colocalization for several variants. Interestingly, two of these findings were associated with incretin measurements during the OGTT. The T risk allele of rs7903146 in *TCF7L2* influenced higher total GLP-1 levels in our study as previously reported,^8^ but here we provide evidence of colocalization to further support the hypothesis that rs7903146 affects T2D risk by modulating incretin action.^8,15,16^ We also demonstrate that the protective C allele of rs703972 near *ZMIZ1* was associated with increased levels of active GLP-1. Interestingly, *ZMIZ1* has been previously reported to play a role in regulation of β-cell function, with expression of *ZMIZ1* reducing insulin secretion.^28^ Thus, an augmented incretin response may explain how C-allele carriers are able to mitigate their T2D risk. Incretins have been implicated in the pathophysiology of T2D; however, it is unknown whether the incretin effect is impaired due to a reduction in functional β-cell mass or due to a defect in incretin action leading to resistance.^29^ Our colocalization analysis provides support for altered incretin physiology in the pathogenesis of T2D and sheds additional insight on a potential mechanism underlying the effect of the *ZMIZ1* variant.

Previously, we reported that a higher T2D polygenic score of 65 variants was associated with several measures of glipizide response at nominal significance,^24^ but we did not identify any associations with phenotypes of metformin response. With the availability of genome-wide genotyping and access to full summary statistics from larger meta-analyses for T2D and glycaemic traits, we expanded our analysis to incorporate large numbers of sub-significant variants across the genome. With a T2D gePS, we confirmed the previous association between a higher genetic burden for T2D and greater glucose AOC, indicating an enhanced response to glipizide. Moreover, we found that individuals with a greater burden of risk variants for higher fasting glucose, possibly representing a genetic susceptibility for lower β-cell function, had a diminished response to metformin. Our ability to detect this pharmacogenetic association was likely bolstered by the vast increase in the number of variants included in the polygenic score and may have clinical implications for the effectiveness of metformin as a first-line therapy in those genetically predisposed to fasting dysglycaemia. While we hypothesized that physiologically-derived clusters related to the drug’s mode of action may have an influence on the acute drug response (i.e., association between β-cell function clusters and glipizide response), we did not detect associations with primary outcomes of metformin or glipizide response, possibly due to lower statistical power of the pPS, which comprise a smaller number of variants. Given the increasing availability of genotype information, future studies are needed to validate the utility and predictive value of polygenic scores for drug response.

Our study is the first GWAS of acute metformin and glipizide response including participants at risk of T2D from multiple ancestries. In contrast to existing T2D pharmacogenetic GWAS performed in European populations,^6,7,12^ over a third of SUGAR-MGH participants were of non-European descent. The value of analysing cohorts that span multiple ancestries is exemplified by the identification of novel associations in genetic variants that are more prevalent in non-European populations. Several of our genome-wide significant findings (rs149403252 near *ERC2*, rs9954585 near *TXNL1*, and rs111770298 near *BABAM2*) had minor allele frequencies that were common to low-frequency in African populations and rare in European populations. Associations near these genes have not previously been identified as related to T2D risk or response to anti-diabetes medications, which may be due to the dearth of studies in non-European populations. Understanding the impact of such ancestry-specific variants may guide treatment decisions for T2D in these population subgroups in the future, but also provide drug targets suitable for all ancestries. One major barrier to translate ancestry-specific variants to their function, is the lack of ancestry-specific genetic and genomic data. For instance, the Genotype-Tissue Expression (GTEx) project largely contains individuals with European ancestry,^30^ limiting our ability to characterize the effects of genome-wide significant variants not present or at low frequency in Europeans on the transcriptome across human tissues. Similarly, the lack of phenome-wide association data on diverse ancestries hinders follow-up of identified variants. Expansion of existing datasets to include non-European populations will be valuable for linking pharmacogenetic associations to functional mechanisms.

Given the global dearth of pharmacogenomic GWAS, especially those conducted in non-European populations, one major limitation of this work is the absence of an adequate replication venue. Due to the unique characteristics of this study, no comparable replication venue was readily available. Nonetheless, we believe that our hypothesis-generating findings could spearhead additional data generation in future cohorts. Another limitation is that the study design did not incorporate a baseline OGTT, which limited our ability to assess the impact of metformin on a dynamic glucose challenge. This was due to the financial and time constraints of enrolling subjects for an additional OGTT. A final limitation is that our sample size was small for measurements of incretin levels, which restricted our ability to detect additional findings relevant to incretin physiology.

In summary, we identified novel genetic variation in a multi-ethnic human drug perturbation study that requires validation in ancestry-specific cohorts but has the potential to influence the selection of anti-diabetes medications in specific populations. We demonstrated the utility of our pharmacogenetic resource for understanding the underlying mechanisms of known genetic variation for T2D and glycaemic traits. Beyond the primary drug endpoints, we created a public resource to permit the organization and sharing of genetic association results across a wide variety of traits in SUGAR-MGH, which can be used as a validation cohort for future pharmacogenetic discoveries by others as well as functional characterization of newly-identified genes implicated in the pathogenesis of T2D.

## Data Availability

The complete summary statistics from this study will be deposited and made available at the Common Metabolic Diseases Knowledge Portal (https://hugeamp.org) and the GWAS Catalog following final publication in a peer-reviewed journal. Additional data requests should be sent by email to the corresponding author.

## Contributors

All authors took part in designing the experiments presented in this manuscript. VK, LNB, MSU, AL, and JCF recruited participants in SUGAR-MGH. VK supervised patient recruitment, data collection, IRB review and approval, performed DNA extractions, and managed GWAS genotyping. Quality control, imputation of the genetic data, and GWAS analyses were performed by JMM. JHL, LNB, VK, KF, and JMM performed follow-up of GWAS data analysis. JHL, LNB, VK, JMM, and JCF contributed to the interpretation of the results. JHL, LNB, VK, and JMM wrote and prepared the manuscript. All authors were involved in reading and editing the manuscript. JMM and JCF jointly supervised this study. JCF is the guarantor of this work.

## Data sharing

The complete summary statistics from this study will be deposited and made available at the Common Metabolic Diseases Knowledge Portal (https://hugeamp.org) and the GWAS Catalogue following article publication in a peer-reviewed journal. Related study documents, including the original study protocol and informed consent forms are available.^17^ Additional data requests should be sent by email to the corresponding author.

## Acknowledgments

We would like to thank Juan Del Rio for his support with Figure 1 generation. This work was conducted with support from National Institutes of Health/NIDDK awards R01 DK088214, R03 DK077675, and P30 DK036836; from the Joslin Clinical Research Center from its philanthropic donors; and the Harvard Catalyst: The Harvard Clinical and Translational Science Center (National Center for Research Resources and the National Center for Advancing Translational Sciences, NIH Awards M01-RR-01066, 1 UL1 RR025758-04 and 8UL1TR000170-05 and financial contributions from Harvard University and its affiliated academic health care centres). JHL received individual support from NIH T32DK007028. LNB is supported by NIH K23DK125839. MSU is supported by NIDDK K23DK114551. AL is supported by grant 2020096 from the Doris Duke Charitable Foundation. Portions of this study were previously presented as an oral presentation at the 81st Virtual Scientific Sessions of the American Diabetes Association, 25-29 June 2021.

## RESEARCH IN CONTEXT

### Evidence before the study

Treatment of type 2 diabetes (T2D) is currently algorithmic and does not take into consideration the underlying genetics of the individual or the specific disease pathophysiology that might benefit from a tailored intervention. Genome-wide association studies (GWAS) have uncovered genetic loci influencing metformin and sulfonylurea response but were largely performed in European populations with established disease. In the Study to Understand the Genetics of the Acute Response to Metformin and Glipizide in Humans (SUGAR-MGH), we utilized a genome-wide approach to identify new pharmacogenetic associations in a multi-ethnic cohort at increased risk of T2D. Using this resource, we also tested the influence of known genetic risk factors for T2D and related glycaemic traits on SUGAR-MGH outcomes to understand the potential functional relevance of these loci.

### Added value of the study

We identified novel genomic regions that are associated with acute metformin and glipizide response at genome-wide significance. Several of our top findings were more common in African-ancestry individuals, underscoring the importance of studying non-European populations. We also found that established T2D and glycaemic trait loci were associated with differences in incretin levels in SUGAR-MGH with high posterior probabilities of colocalization; these findings provide further insight into incretin physiology as a potential mechanism by which these variants influence T2D risk.

### Implications of all the available evidence

Taken with a view to advance precision medicine, our study provides initial evidence for the consideration of ancestry-specific variants on the choice of pharmacotherapy for T2D. Furthermore, we present a richly phenotyped pharmacogenetic resource that can contribute valuable information about the potential mechanisms of action of T2D-related genetic variation. The public sharing of our genetic association results from this study will enable others to search or validate newly identified genetic loci related to T2D or response to T2D medications.

